# Cell type-specific methylome-wide association studies of childhood ADHD symptoms

**DOI:** 10.1101/2025.04.16.25325956

**Authors:** Mandy Meijer, Marieke Klein, Doretta Caramaschi, Shaunna L. Clark, Marta Cosin-Tomas, Nastassja Koen, Xueling Lu, Rosa H. Mulder, Stefan W. Röder, Yining Zhang, Lea Zilich, Mariona Bustamente, Michael Deuschle, Janine F. Felix, Juan Ramos González, Regina Gražulevičiene, Fabian Streit, John Wright, Angel Carracedo, Charlotte A.M. Cecil, Eva Corpeleijn, Catharina A. Hartman, Gunda Herberth, Anke Huels, Caroline Relton, Harold Snieder, Dan J. Stein, Jordi Sunyer, Stephanie H. Witt, Heather J. Zar, Ana C. Zenclussen, Barbara Franke, William Copeland, Karolina A. Aberg, Edwin J.C.G. van den Oord

## Abstract

**Objective:** Studying DNA methylation (DNAm) can provide insights into gene-regulatory mechanisms underlying attention-deficit/hyperactivity disorder (ADHD). While most DNAm studies were performed in bulk tissue, this study used statistical deconvolution to identify cell type-specific DNAm profiles, from five major blood cell types, associated with childhood ADHD symptoms.

**Methods:** We performed meta-analyses of methylome-wide association studies (MWAS) for ADHD symptoms (age_range_=4-16 years) in peripheral blood collected during childhood and in cord blood. The investigated cohorts included seven array-based methylation datasets assaying up to 450K CpGs from the Pregnancy And Childhood Epigenetics Consortium (N=2 934 peripheral blood; N=2 546 cord blood) and a sequencing-based methylation dataset assaying nearly all 28 million CpGs in blood from the Great Smoky Mountain Study (GSMS; N=583).

**Results:** The meta-analyses resulted in methylome-wide significant (FDR<0.05) ADHD associations in CD8T cells (*RPL31P11* and *KCNJ5)* for peripheral blood, and, in cord blood, in monocytes (*PDE6B*), CD8T cells (*KCNA3* and *HAND2*), and NK cells (*KIFC1*). Notably, several significant sites detected in peripheral blood (*RPL31P11* and *KCNJ5*) were also detected in cord blood. Furthermore, extended MWAS of all sites available for GSMS detected 69 and 17 additional CpGs in monocytes and granulocytes, respectively. In this first cell type-specific MWAS for ADHD, we identified DNAm associations for ADHD symptoms; some associations were seen in both peripheral blood and cord blood, suggesting potential susceptibility markers for increased ADHD risk.

**Conclusions:** These findings show that cell type-specific analyses and sequencing-based approaches can increase insights into the epigenetic patterns associated with ADHD symptoms in childhood.

## Introduction

Attention-deficit/hyperactivity disorder (ADHD) is a neurodevelopmental disorder characterized by symptoms of inattention and/or hyperactivity and impulsivity, with a prevalence of 5% in children (1). ADHD symptoms are not only present in individuals with a clinical diagnosis, but are present as a continuous trait in the general population, sharing biological mechanisms with the clinical diagnosis (2, 3). ADHD is a multifactorial disorder to which genetic and environmental factors contribute (3–6). These factors can alter DNA methylation (DNAm) (7), which is an epigenetic modification involved in gene regulation, with potential long-term effects on downstream phenotypes (8). Performing methylome-wide association studies (MWAS; also referred to as epigenome-wide association studies) is a promising approach to study whether DNAm is associated with the development of ADHD, ADHD severity, and changes in ADHD phenotype over time. Furthermore, DNAm studies may yield novel biomarkers for early risk detection, which could facilitate preventive care and intervention (9).

MWAS for ADHD case-control status (10, 11) and ADHD symptoms in single cohorts (11, 12) and larger meta-analytic (13, 14) approaches have been performed for DNAm in whole blood and saliva. For example, DNAm annotated to the genes *SKI*, *ZNF544*, *ST3GAL4*, and *PEX2* in cord blood at birth were prospectively associated with childhood ADHD symptom trajectories (12), and *CREB5*, *ZBTB38*, *PPIL2*, *TRERF1*, and *ERC2* with ADHD childhood symptoms (14). However, these DNAm associations were not observed when examining peripheral blood collected cross-sectionally during childhood (12, 14).

So far, all larger epigenetic studies on ADHD have been performed in bulk tissue, such as whole blood, which contains a mixture of white cell types (15). However, DNAm profiles are cell type-specific (16), and it is well-known that white cell types differ in their roles in functioning and regulation of the immune system as well as in their access to and functions in the developing and mature brain (e.g. 17, 18, 19). In studies of bulk tissue, ADHD-associated DNAm signals that occur in only a single cell type might go undetected, and associations with effects in different directions in different cell types may cancel one another out (20). The fact that both epidemiological and and genetic studies have shown co-occurrence of ADHD and immune-related disorders (3, 21, 22) could make a peripheral blood cell type-specific MWAS for ADHD (symptoms) even more interesting. Also in earlier studies in other phenotypes that identified cell type-specific DNAm profiles, such findings have contributed insight into the underlying biological mechanisms of the studied phenotypes (20, 23–25).

ADHD-related DNAm profiles in peripheral blood collected in childhood or adulthood require different interpretations from those observed in neonatal samples, e.g., in cord blood. DNAm associations with ADHD symptoms observed in cross-sectional studies of peripheral blood cells collected in childhood or adulthood might reflect a mixture of causes and consequences of these symptoms or their correlates (e.g., medication use). In contrast, DNAm associations with ADHD symptoms observed in cord blood, which is collected right after birth and prior to any measurable symptoms, may reflect epigenetic programming *in utero* due to the presence of prenatal and genetic risk factors; this could give clues for predicting a sensitivity to developing ADHD symptoms during childhood.

In previous ADHD-related MWAS (10–14), commercially available arrays have been used containing up to 850 000 DNAm (CpG) sites; this corresponds to approximately 3% of the over 28 million sites that can be methylated in human blood samples. Since the commercial arrays were not designed specifically to investigate DNAm signatures of behavioral traits, the discovery of important DNAm signals might be hampered by the limited coverage of the arrays. Cost-effective analysis methods for a better coverage of the DNA methylome include sequencing approaches, such as methyl-domain-binding sequencing (MBD-seq), which covers nearly all methylation sites in human blood cells (26). Combining DNAm assessments measured using different technologies can increase the robustness of DNAm associations by eliminating possible false discoveries caused by assay-specific technical artefacts.

Here, for the first time, we investigated cell type-specific DNAm patterns for childhood ADHD symptoms in both, peripheral blood collected during childhood (N=2 934 from 5 cohorts) and neonatal (i.e., cord) blood (N=2 546 from 7 cohorts); using data from the PACE consortium, we applied a well-established statistical deconvolution to convert bulk DNAm data to cell type-specific signals (15, 20, 27). We compared findings from peripheral and cord blood to identify time-point-specific and consistently ADHD-associated DNAm sites. In addition to commercial array-based analysis of up to 450 000 CpG sites, we were able to also investigate data from DNAm sequencing of nearly all 28 million CpG sites available in a subset of samples (N=583 children, age_range_=9-16 years) enabling us to also explore methylation sites not covered by standard arrays.

## Methods and materials

This section provides a concise description of the methods. Additional details can be found in the supplemental material that is referred to here by using prefix S (e.g., S1.1 refers to section 1.1 in the supplemental material). A schematic overview of our study can be found in Figure 1.

**Figure 1.**
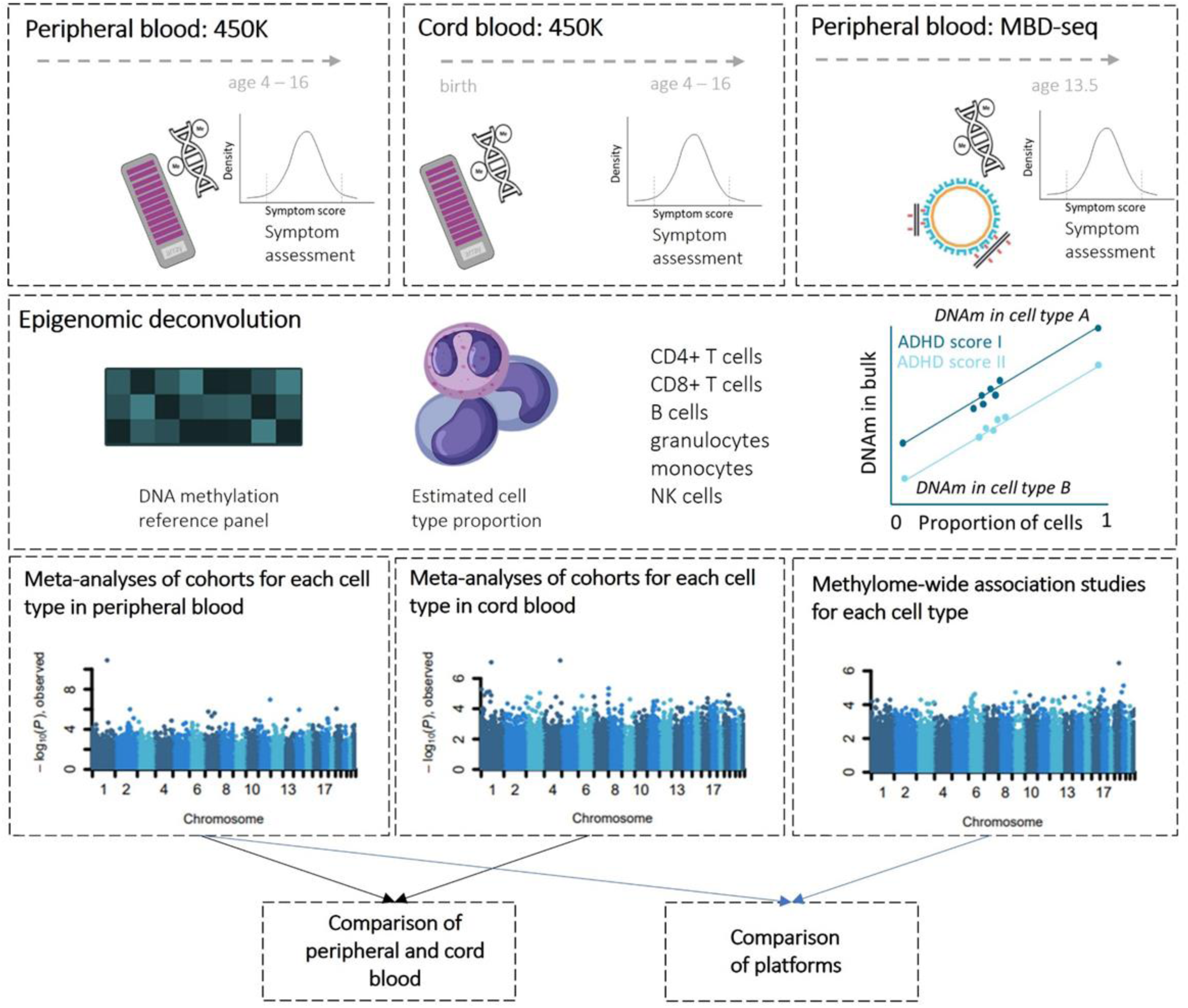
Study overview. Top row: General population cohorts of the PACE consortium and the GSMS cohort were included in the current study. Peripheral blood was collected at age 4-10 years (450K; left) and age 9-16 years (MBD-sequencing, right). Cord blood was collected at birth (middle). ADHD symptom scores were assessed at the same time peripheral blood was collected. DNAm was measured with the 450K array covering up to 450 000 DNAm sites in the genome (left, middle) or MBD-sequencing (right), covering nearly all 28 million CpG sites in the human genome. Second row: Epigenomic deconvolution of DNAm data was performed by estimating cell type proportions based on a DNAm reference panel; cell type-specific DNAm values were extrapolated from bulk data based on cell type proportions. Third row: Cell type-specific meta-analysis of overlapping sites from MWAS in peripheral (including sites retrieved from MBD-seq covered by the 450K array) and cord blood for the CpG sites covered on the commercial arrays, and a cell type-specific MWAS in peripheral blood from GSMS (right) of nearly all 28 million CpGs. Bottom row: Comparisons of associations over time were made by comparing identified sites from the meta-analysis of peripheral blood with the results from the meta-analysis of cord blood, and vice versa. Similarly, we performed comparison between the 450K array and MBD-seq platforms.

### Cohorts and measures

Eight cohorts from the PACE consortium (ALSPAC, (28, 29), Generation R Study (GenR, (30)), INMA (31), HELIX (32), DCHS (33), GECKO (34), LiNA (35), POSEIDON (36), and GSMS (37)) were included in the meta-analyses, see information on the individual cohorts in S1.1. In all cohorts, ADHD symptoms were assessed during childhood (age 4-16 years) with either the Development and Well-being Assessment (DAWBA) (38), Child Behavior Checklist (CBCL) (39), Strengths and Difficulties Questionnaire (SDQ) (40) or Parent and Child Interview-based Structured Diagnostic Interview (PACA) (37) (see S1.1). Given the usage of different instruments to assess ADHD symptoms, total ADHD symptom scores were standardized by scaling measures within cohorts to have a mean of zero and standard deviation of one, as previously described for a meta-analysis of bulk tissue of a subset of the cohorts (14). Cord blood was collected at birth. Peripheral blood was collected at the same time childhood ADHD symptoms were collected. Procedures were approved by the relevant ethics committees for each cohort, and all parents provided informed consent for participation of their children.

### Methylation assays

Methylation was assessed with either the Illumina Infinium HumanMethylation450 BeadChip (450K) (41) or an optimized protocol for methyl-CG binding domain sequencing (MBD-seq (42); GSMS cohort).

Elsewhere we summarized key features of MBD-seq using empirical data (43). Critically, MBD-seq achieves near complete coverage of all 28 million sites in the blood methylome (44, 45). For all cohorts with 450K array data, quality control of samples and CpGs was performed using the same pipeline (S.1.2.1, see (46) for further details). For the MBD-seq data, we performed quality control of samples, reads, and CpG sites (S.1.2.2, further details in (47)) using the RaMWAS package (48).

Similar to other deconvolution approaches (20, 49–52), we used statistical epigenomic deconvolution to perform cell type MWASs from bulk methylation data. Deconvolution was first introduced over 20 years ago (49). It is commonly used in gene expression studies (49, 50) and can readily be applied to methylation data (20, 51, 52). The approach has been validated by showing it can detect known associations in artificial data as well as associations observed with empirical data from purified cells (20, 50). S1.3 provides an intuitive and a formal description of the method. In summary, cell type proportions were first estimated for each bulk sample individually using a reference panel (15, 53). Reference panels are comprised of methylation profiles of physically sorted subtypes of cells generated from a limited number of subjects. The estimated cell type proportions were then used to test for cell type-specific associations with the RaMWAS R package in every individual cohort and for cord and peripheral blood separately.

To ensure optimal estimates of cell type proportions, the reference panels were specific for each sample type and data type. For 450K array data, cell types in cord and peripheral blood included CD8^+^ T-cells (CD8T), CD4^+^ T-cells (CD4T), CD14^+^ monocytes, CD15^+^ granulocytes, CD19^+^ B-cells, and CD56^+^ natural killer cells (NK), based on the *FlowSorted.Blood.450K* R package. For cord blood, the reference panel also included nucleated red blood cells, but we did not perform cell type-specific MWAS on these cells, as they are not present in peripheral blood and thus will not support comparisons between cord and peripheral blood. For MBD-seq data, we used a MBD-specific reference panel (54, 55) that comprised CD3^+^ T-cells (which include both CD8^+^ and CD4^+^ T cells), CD14^+^ monocytes, CD15^+^ granulocytes, and CD19^+^ B-cells.

In the linear regression models of the MWASs, we controlled for potential (cohort-specific) confounders by regressing out demographic and clinical covariates (a.o., sex, age, and cell type composition) (Table S1). In addition, we regressed out a series of technical covariates derived from the methylation data (S1.2). Finally, we performed a principal component analysis on the methylation data after regressing out all measured covariates to capture any remaining unmeasured covariates. The main principal component from these analyses was regressed out to account for any remaining unmeasured sources of confounding in every single dataset.

### Meta-analyses

A fixed-effects meta-analysis of all cohorts was performed with METAL with the Stouffer method (56). Heterogeneity was calculated to determine the statistical heterogeneity between studies by weighting the deviation of each study’s observed effect from the summary effect by the inverse of the study’s variance, where a low p-value from the chi-squared test indicates high heterogeneity.

As the MBD-seq reference panel did not include CD4^+^ T and CD8^+^ T cells, results for CD3^+^ T cells (which includes CD4^+^ T and CD8^+^ T cells) from MBD-seq data were included in the analyses for both CD4^+^ T cells and CD8^+^ T cells. To analyse array and MBD-seq data together, we used the 230 231 CpG sites from the MBD-seq that we were able to match to any of the ∼450 000 CpG sites covered on the array before quality control (S1.2).

To account for multiple testing, we controlled the false discovery rate (FDR) (57) for each analysis at the 0.05 level. We chose this method, since it allowed us to keep the expected proportions of false discoveries among the significant findings at 5% regardless of whether array or MBD-seq data was analysed; this would not have been possible with methods that control the family-wise error rate, such as the Bonferroni correction (58, 59).

### Reactome and gene ontology analyses

To study pathways and biological functions, enrichment analyses were performed using the Reactome (60) and Gene Ontology (GO) databases (61) (S1.4). We ran separate analyses for “suggestive” association findings (q-value<0.5) for cord blood and peripheral blood meta-analyses and for the MWAS performed on the GSMS cohort only, all including a 100 kb flanking region for the genes. We used circular permutations to properly control the Type I error in the presence of correlated sites (see (62) and Figure S1.4.1). Furthermore, as the permutations were performed on a CpG site level, they also properly account for gene size, as genes with more CpG sites are more likely to be among the top results in the permutations.

### Comparison of data sources

To investigate whether the identified DNAm associations replicated across tissue and time, we looked up the association strength of the significant findings from peripheral blood in cord blood, and vice versa. Look-up associations were considered significant after Bonferroni correction for multiple testing allowing for a type I error of 0.05. We determined whether the number of significant look-up findings was higher than expected by chance by comparing the actual number of significant findings to the null probability of finding sites significant with an FDR-adjusted p-value<0.05 in both meta-analysis of peripheral and cord blood. This comparison was done with a 1-sample proportions test with continuity corrections, without controlling for correlated DNA methylation sites in the genome.

## Results

### Cohort descriptions

The total sample size of the cohorts investigated was 2 546 for cord blood and 2 934 for peripheral blood (Table 1). After quality control, the total number of autosomal CpG sites for the array studies was 374 392 for peripheral blood and 375 362 for cord blood. Akin to filtering SNPs with low minor allele frequency, we excluded 4 881 020 CpG sites from the MBD-seq data, as they were rarely methylated. This left 22 670 747 autosomal CpG sites for analysis.

**Table 1.**
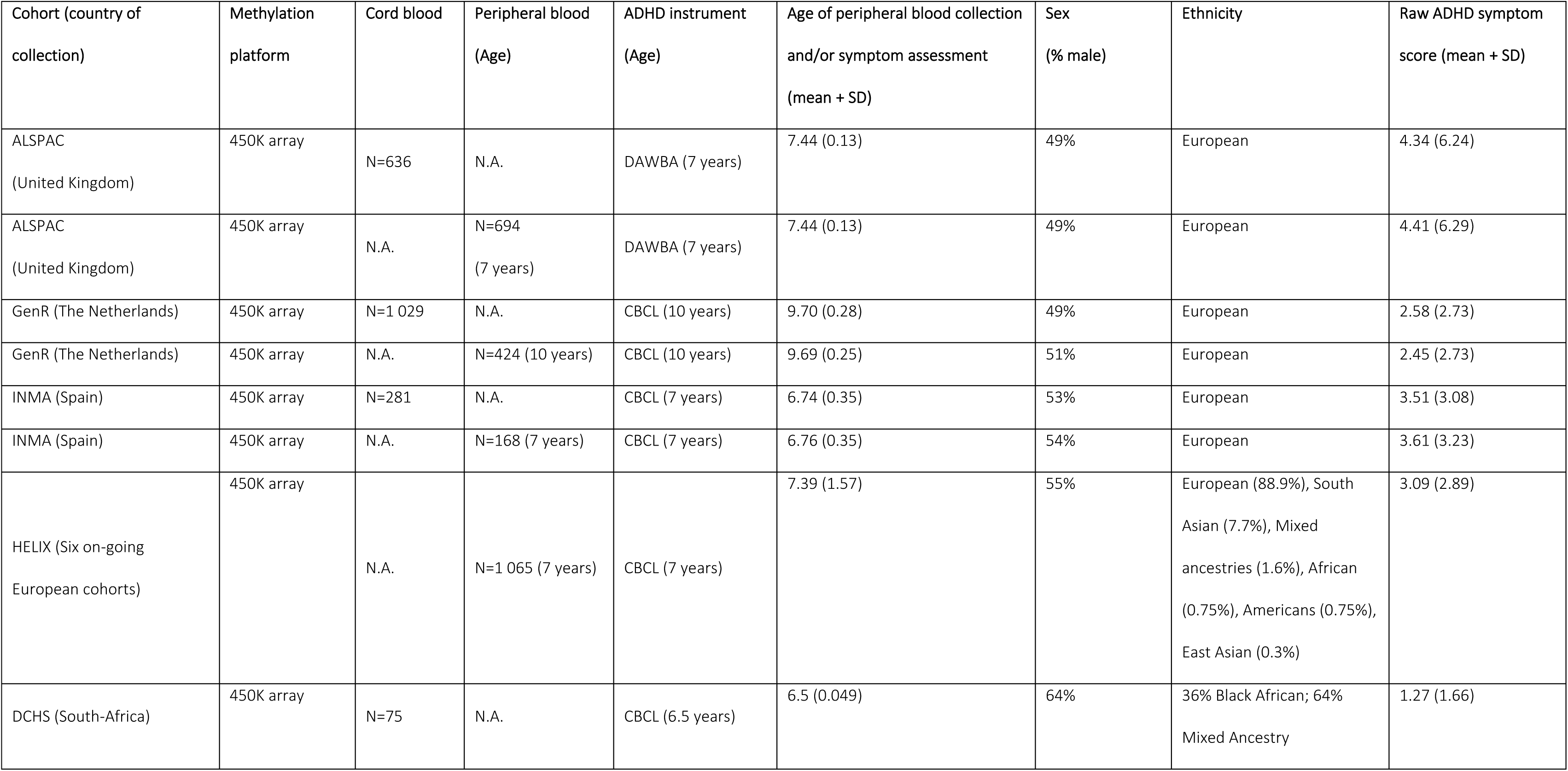
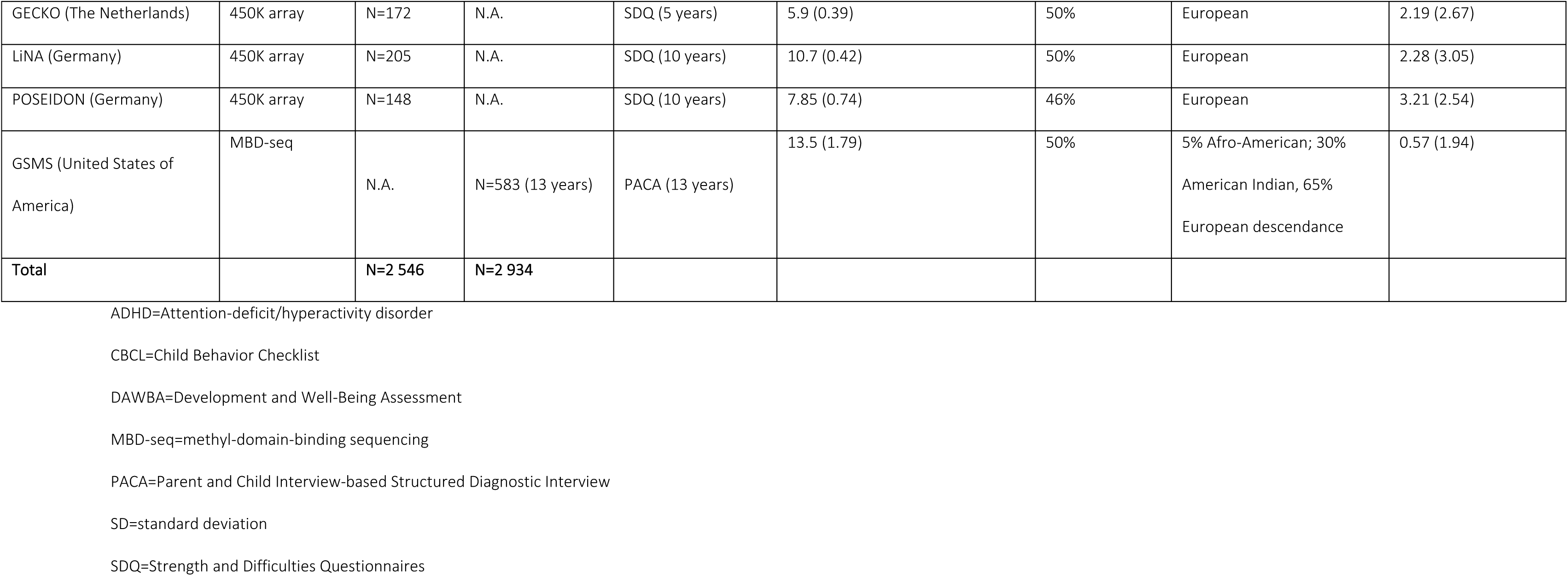
Demographics of cohorts.

### Cell type-specific DNAm meta-analyses

We aimed to identify cell type-specific DNAm associations with childhood ADHD symptoms assessed at age 4-16 years in peripheral blood obtained at the same time as symptom assessment (cross-sectional MWASs). Here, we focused on all DNAm sites covered by the 450K array after QC and overlapping with the MBD-seq data (230 231 CpG sites). Meta-analyses of the individual MWASs (meta-MWASs) for individual cell types showed no evidence of inflation or deflation of test statistics (lambda meta-analyses 0.95 to 1.03) (Figure 2A, Figure S2.1). Two DNAm sites, located in Ribosomal Protein L31 Pseudogene 11 (*RPL31P11*) and Potassium Inwardly Rectifying Channel Subfamily J Member 5 (*KCNJ5*) (Table 2), reached a methylome-wide significant association with ADHD symptoms in CD8T cells. Both sites showed low heterogeneity p-values (1.00*10^-3^<p-value<0.02), indicating heterogeneity between cohorts. The significant DNAm sites in CD3T cells in the GSMS cohort - assessed with MBD-seq rather than the 450K array - showed the same direction of effect in the meta-analysis for CD8T cells (Table 2, Table S2.1), showing cross-method robustness. When removing the GSMS cohort, and potentially the effects of CD4T cells also captured by CD3T cell proportions, both sites remained significant (1:161655124, Z-score=7.875, p=3.41*10^-15^; 11:128781826, Z-score=-5.614; 1.98*10^-8^). Furthermore, none of the results were driven by ethnicity (Table S2.3).

**Figure 2.**
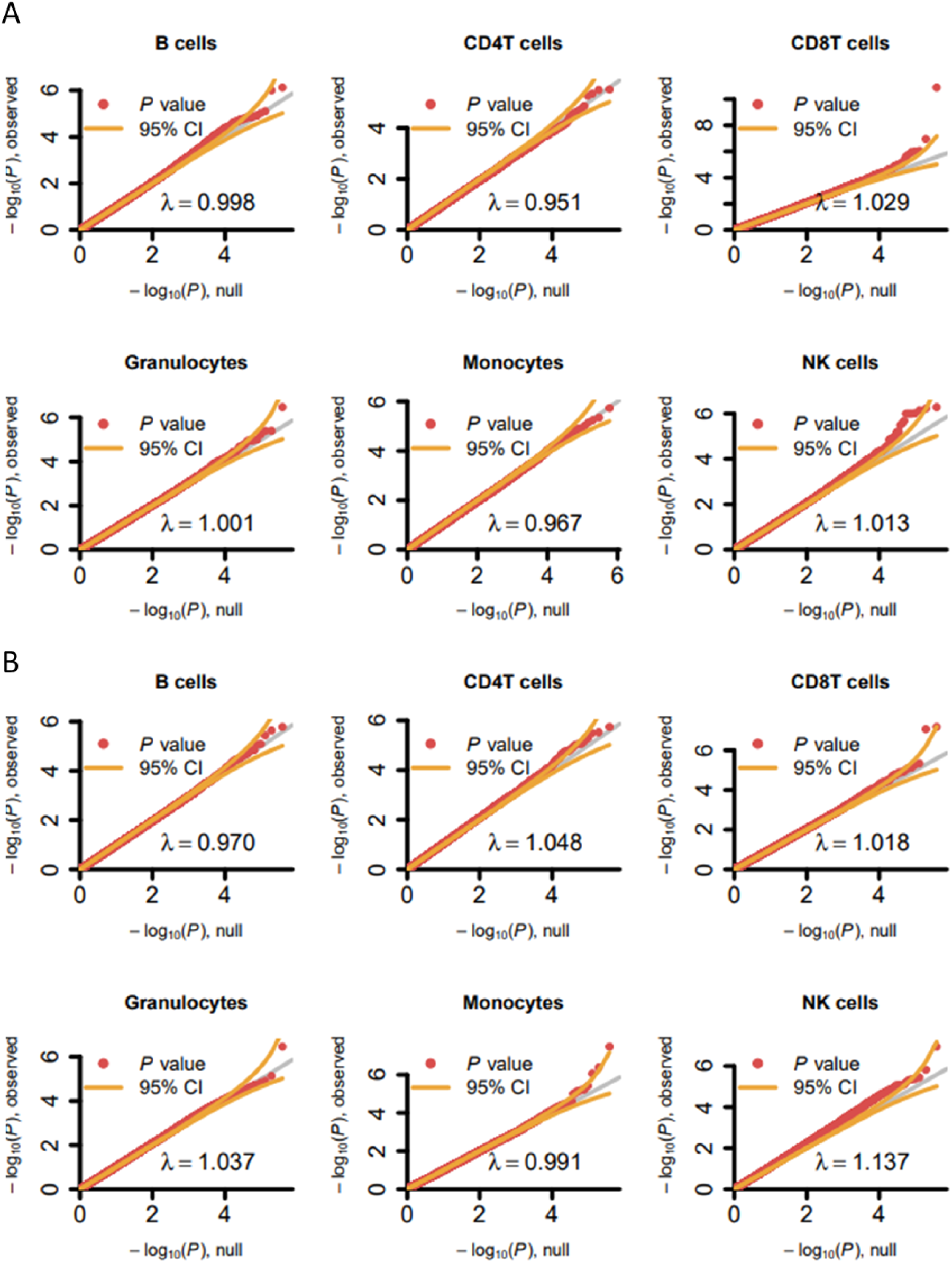
Quantile-quantile (QQ) plots for methylome-wide association meta-analyses of array-based DNA methylation sites for each cell type in peripheral and cord blood. A) QQ plots for meta-analysis of DNAm in peripheral blood and symptom scores collected in childhood. Peripheral blood includes DNA methylation data from the GSMS, except for NK cells, since this data was not available. B) QQ plots for meta-analysis of DNAm in cord blood at birth analyzed with symptom scores collected later in childhood. The x-axis shows the expected-log_10_(p-value) and the y-axis shows the observed-log_10_(p-value). The inflation factor (λ) is given at the bottom of each plot. Orange lines indicate the 95% confidence interval.

**Table 2.**
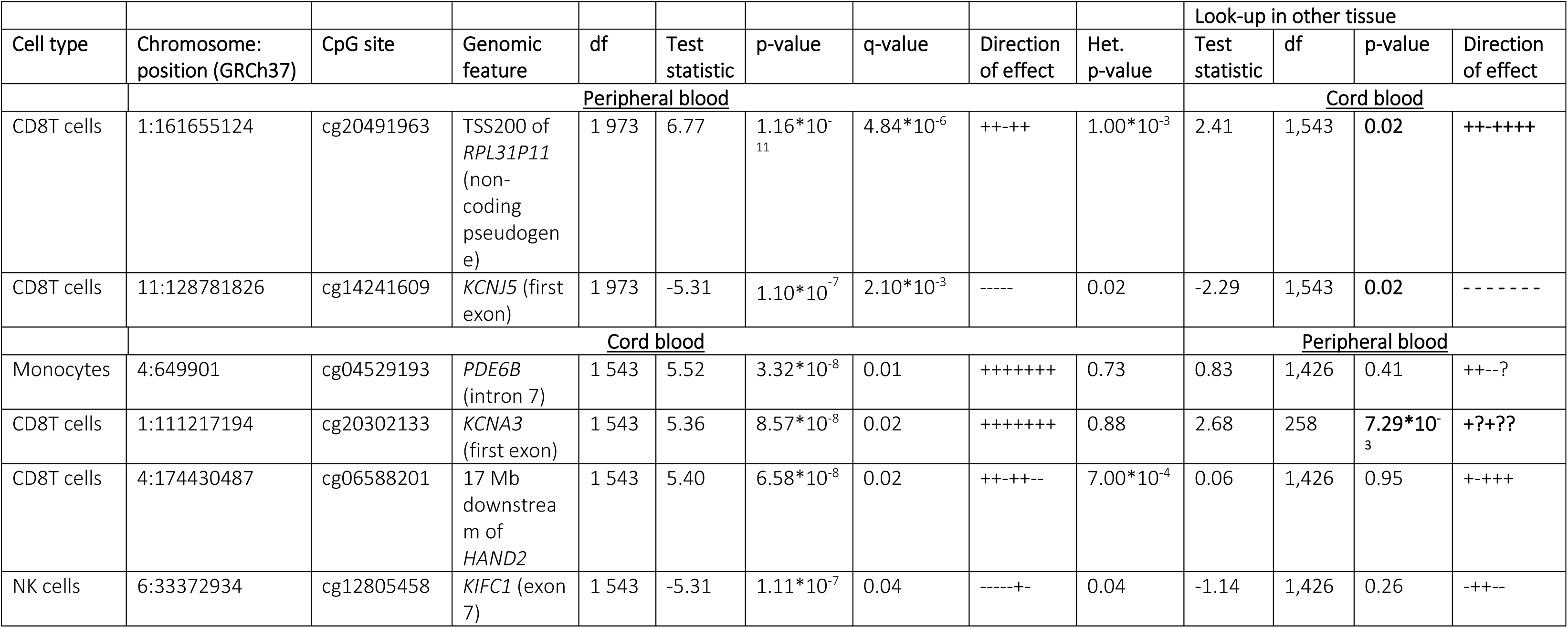
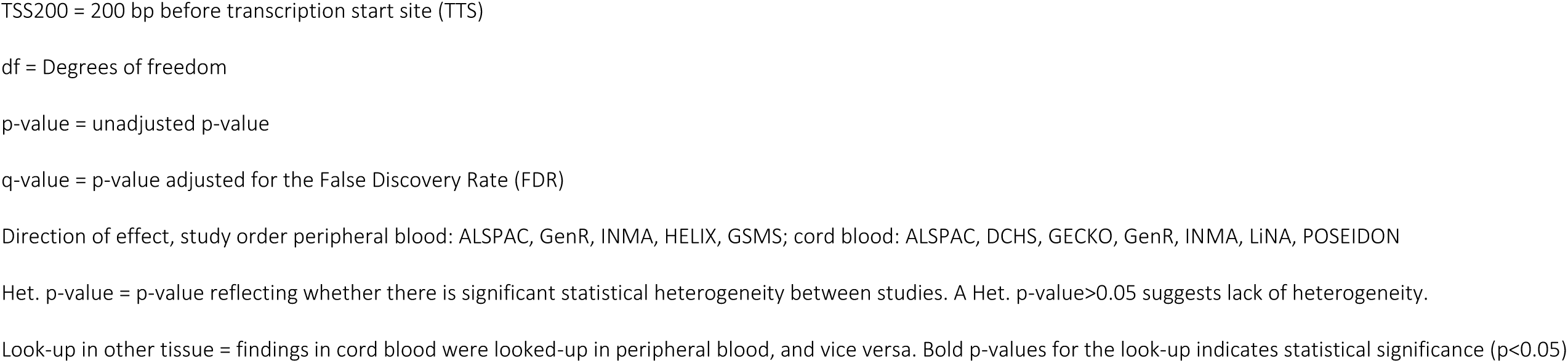
Methylome-wide significant findings from cell type-specific MWAS meta-analysis on ADHD childhood symptoms in cord-and peripheral blood.

Like for peripheral blood, neither inflation nor deflation of test statistics was detected for the meta-MWASs of cord blood (lambda meta-analyses 0.97 to 1.17) (Figure 2B). In cord blood, we identified methylome-wide FDR-adjusted (p<0.05) significant findings in monocytes (annotated to *PDE6B*), CD8T cells (annotated to *KCNA3* and *HAND2*), and NK cells (annotated to *KIFC1*) (Table 2). Especially the findings in Phosphodiesterase (*PDE6B*) and Potassium Voltage-gated Channel Subfamily A Member 3 (*KCNA3*) showed low study statistic heterogeneity.

To investigate whether the identified DNAm associations with ADHD symptoms were present across two time points, we looked-up the identified sites from peripheral blood meta-MWASs in the meta-MWASs in cord blood, and vice versa. Of the two DNAm sites identified in CD8T cells in peripheral blood, both were Bonferroni-significant in CD8T cells in cord blood. These sites were annotated to *RPL31P11* (Bonferroni-corrected p-value=0.02), and *KCNJ5* (Bonferroni-corrected p-value=0.02). Of the four DNAm sites identified in cord blood, one, annotated to *KCNA3*, also appeared significantly associated with ADHD symptoms in peripheral blood CD8T cells (Bonferroni-corrected p-value=7.29*10^-3^; Table 2); however, only two of the five samples had this site available for analysis.

### High resolution cell type-specific MWASs identify additional DNAm sites associated with ADHD symptoms

The commercial arrays leave the majority of DNAm sites in the human genome uncovered. Therefore, in addition to our main meta-analyses, we performed MWASs of ADHD symptoms using the full coverage provided by the MBD-seq data available in the GSMS cohort (including 22 670 747 DNAm sites after quality control). Test statistics did not indicate inflation or deflation (MWAS lambda range: 0.87 – 1.00) in most cell types except for B cells, where the lambda was 0.68 (Figure 3), probably driven by the low estimated B cell proportions. We therefore excluded the B cell results from this analysis. Across the cell type-specific MWASs, we identified 69 methylome-wide significant associations (q-value<0.05) implicating 21 genomic loci in monocytes (CD14^+^ cells), and 17 associations implicating eight genomic loci in granulocytes (CD15^+^ cells) (Figure 3, Table S2.2, Figure S2.2). No methylome-wide significant findings were detected for CD3^+^ T cells (Figure S2.2). The most significant findings annotated to genes were located in Alpha-1,6-Mannosylglycoprotein 6-Beta-N-Acetylglucosaminyltransferase B (*MGAT5B;* q-value=7.77*10^-5^) for monocytes and in Semaphorin-4D (*SEMA4D*; q-value=4.17*10^-8^) for granulocytes. In both cell types, we found the same two CpGs located in Scaffold Protein Involved in DNA Repair (*SPIDR*) to be associated with ADHD symptoms, but with effects in opposite directions. We looked up whether genes associated with ADHD childhood symptoms in previously studies in bulk tissue, were also present in the cell type-specific MWAS (12). We found suggestive significant (p<5*10^-5^) associations in the genes *CREB5* (monocytes and granulocytes) and *ERC2* (monocytes).

**Figure 3.**
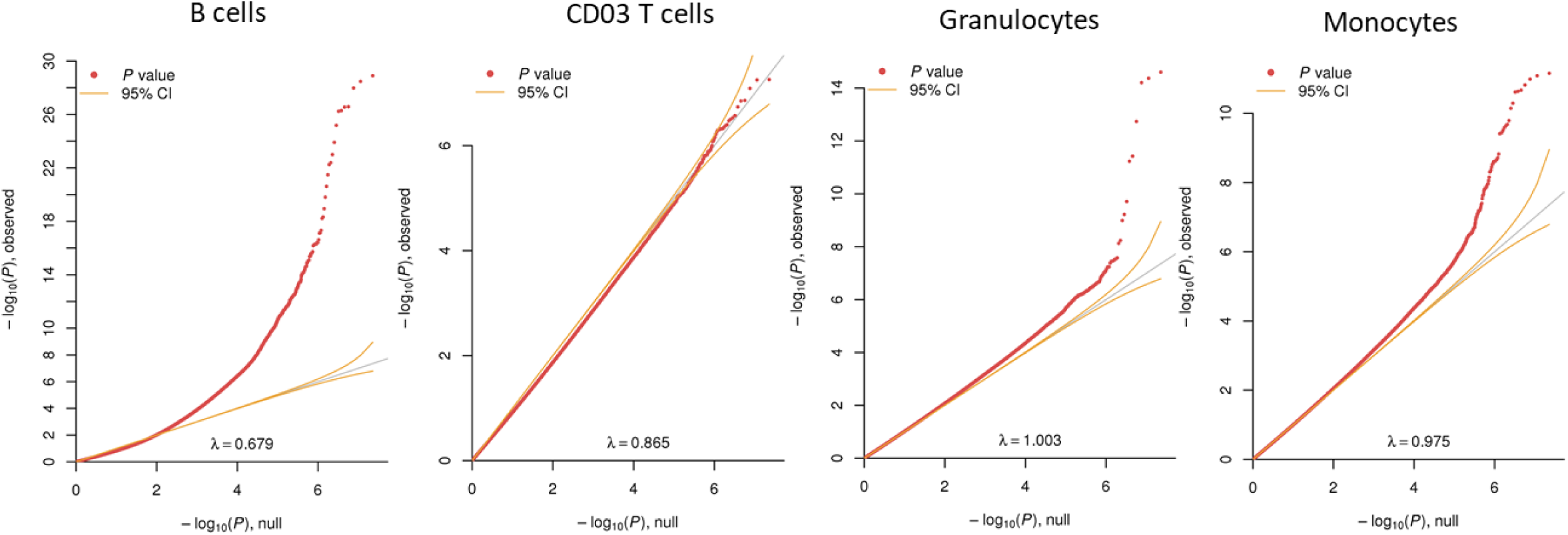
Quantile-quantile plots of cell type-specific MWAS for ADHD symptoms in the GSMS cohort. QQ plots for MWAS of peripheral blood at childhood analyzed with symptom scores collected in childhood. The x-axis shows the expected-log_10_(p-value) and the y-axis shows the observed-log_10_(p-value). The inflation factor (λ) is given at the bottom of each plot. Orange lines indicate the 95% confidence interval.

### Reactome and gene ontology analyses

To identify cell type-specific biological pathways of relevance to ADHD symptoms, we performed Gene Ontology and Reactome enrichment analyses on the results of the cell type-specific meta-MWASs in cord blood and peripheral blood (from the latter, we excluded the GSMS cohort to avoid double dipping), and in the extended MWAS in GSMS. We included all associations for which we detected suggestive significance (q-value<0.5). Significant enrichments (p-value<0.05) were seen for CD8T cells, granulocytes, and NK cells in the meta-MWASs of peripheral blood. These enrichments were mainly related to calcium ion binding and plasma membrane adhesion (Table S2.4). Meta-MWASs in cord blood showed enrichments in monocytes, CD4T cells, and CD8T cells (Table S2.4). Enrichments in the MWASs of the GSMS cohort were observed in granulocytes and monocytes; those were mainly related to the synapse and neurons (Table S2.4).

## Discussion

We performed a meta-analysis of methylome-wide association studies for childhood ADHD symptoms going beyond the current state of the art by performing cell type-specific analyses in both cord blood taken at birth and peripheral blood from later in childhood. This study design offered several unique advantages. First, using DNA methylation data from individuals for which blood samples and phenotypic information were collected at the same time allowed for cross-sectional investigations of associated methylation markers. Second, performing association testing between ADHD symptoms measured in childhood and methylation profiles of neonatal cord blood collected prior the presence of any measurable symptoms, allowed for identification of DNAm sites potentially indicating increased susceptibility for ADHD symptoms. Comparing the results between the neonatal and childhood analyses allowed us to pinpoint those susceptibility markers that are consistently associated with ADHD symptoms over time. Lastly, being generated using two different techniques and across several different cohorts, the methylation data available allowed us to compare results across platforms and cohorts to ensure that significant findings were not driven by platform-specific artifacts or cohort-specific characteristics.

The meta-MWAS of cross-sectional peripheral blood identified two methylome-wide significant associations in CD8T cells. One was located close to *RPL31P11*, a gene without a known function. The second was located in *KCNJ5*. The most important role of the protein encoded by this gene is the regulation of potassium inflow into the cell. Genetic variants in this gene have been associated with ADHD previously, potentially through affecting transcription levels of the gene (63), and full knock-out animal models show learning impairments (64). Our look-up analysis showed that both findings were also associated with ADHD symptoms in neonatal cord blood, suggesting that decreased levels of the methylation marks are present before ADHD symptoms are detectable; given the robustness of those findings across analyses within our study and consistency with genetic findings from earlier studies, these sites may thus be susceptibility markers involved in ADHD symptom development rather than consequences of ADHD-related behavior.

In the cord blood meta-MWASs, we identified four methylome-wide significant associations. This involved associations in *KCNA3* and *HAND2* in CD8T cells, one association located in *PDE6B* in monocytes, and one in *KIFC1* for NK cells. Of those four, the association in *KCNA3* appeared consistent between cord and peripheral blood, but only two of five samples contributed data to this analysis. The *KCNA3* association is interesting for follow-up, as this is the second potassium channel-encoding gene among our findings; the gene is expressed in brain and in T cells, plays a role in neurotransmitter release (65), and its expression is induced by inflammation (65). Cortisol, the main stress hormone, also inhibits the channel encoded by the gene (66). None of the most significant findings were annotated to a known ADHD risk-gene based on genome-wide association studies (3).

Supported by our results, we argue that cell type-specific methylation associations, potentially reflective of susceptibility markers for ADHD symptoms, are present already in the neonatal blood. Some of these methylation associations are stable over time and can, in fact, be detected as highly significant methylation marks in peripheral blood later in childhood. It has previously been noted for several other traits, such as prenatal smoking (67), that epigenetic associations show stability over time. Other highly significant associations detected in neonatal samples did not show association with ADHD symptoms in blood taken in childhood, consistent with the dynamic nature of methylation (67, 68). These factors might be more reflective of environmental risk factors for ADHD symptoms. However, the field needs more and larger longitudinal studies with repeated measures of DNAm and behavior in the same individuals to draw conclusions on stability and origin of DNAm in the context of ADHD.

As the 450K arrays capture only a fraction of all DNAm sites, we took advantage of the much larger number of CpG sites generated with MBD-seq available for the GSMS data set to perform extended cell type-specific MWASs for ADHD symptoms involving over 22 million CpG sites. Studying ∼80% instead of only 2% of CpG sites in the human genome identified 86 additional significant findings. Some of these findings, located in the coding region of *SEMA4D* (granulocytes)*, MGAT5B* (monocytes), and *SPIDR* (granulocytes and monocytes) are of particular interest for ADHD symptoms. SEMA4D is a semaphorin, and an axon guidance cell membrane protein, needed for brain development (69), which interacts with the immune system through B cell activation (70). An integrative analysis of genome-wide association studies found that genes associated with ADHD are related to SEMA4D signaling (71). Alpha-1,6-Mannosylglycoprotein 6-Beta-N-Acetylglucosaminyltransferase B (MGAT5B) is highly expressed in the brain and is involved in the synthesis of surface N-glycans, which are necessary for cell adhesion in brain development (72). Lastly, SPIDR is highly expressed in the cerebellum where it plays a role in DNA double-strand break repair via homologous recombination (72, 73). Genetic variants in *SPIDR* are associated with monocyte count (74). Recently, DNAm of this gene in blood has been associated with neurodegenerative diseases (75).

Our enrichment analyses revealed enrichments related to calcium signaling and the synaptic membrane. Also, on the single gene level we found DNAm associations with ADHD symptoms in genes with roles in brain development and behavior. The discovered DNAm patterns thus suggest brain involvement, in our own study as well as in others on ADHD and related behaviours (10–14), and the question remains why these DNAm profiles are observed in blood, and even in specific blood cell types. Several suggestions have been made, for example that peripheral tissue might carry methylomic signatures resulting from epigenetic programming during embryogenesis (76), and/or that genetic polymorphisms acting on those sites might result in identical methylomic patterns across tissues (77, 78). However, it is also known that specific blood cells, e.g. CD4T cells, are important in brain development (e.g., 17), while others, such as CD8T cells, play a role in brain injury and neurodegeneration (e.g., 79). More generally, inflammatory processes are known to be relevant for ADHD: comorbidity and genetic correlation between immune-related diseases and ADHD has been observed (3, 18), and an inflammatory profile seems to increase the risk for developing ADHD (80, 81). Furthermore, ADHD can occur after traumatic brain injury (e.g. 82), which is known to elicit inflammatory responses. Several of the genes identified for ADHD symptoms in the current study also play roles in inflammation; for example, calcium signaling disruption in CD8T cells leads to auto-immune disorders (83). Also, in previous MWAS for ADHD, evidence for DNAm associations with ADHD in genes related to immune functioning has been observed (11, 13).

Did the extra effort of performing the meta-MWASs in our study in a cell type-specific manner rather than analyzing the data traditionally in bulk pay off? We would argue that this is indeed the case for several reasons. Firstly, many associations were only seen in single cell types, for example, we found the associations with ADHD symptoms to be concentrated in particular in the CD8Tcells (in the meta-analytic part of our study), which play important roles in brain injury and neurodegeneration (79). Such associations would have likely been missed in bulk analyses. Secondly, the methylation sites in the *SPIDR* gene were associated with ADHD symptoms in monocytes and granulocytes, but with opposite directions of effect. These associations would have cancelled each other out in bulk tissue analyses. Thirdly, more DNAm associations and larger effect sizes compared to previous studies on childhood ADHD symptoms performed in bulk were found (14).

Our study should be viewed in the context of its strengths and limitations. Clear strengths are the relatively large sample sizes, the harmonized QC pipelines, the possibility of studying cell type-specific DNAm association with ADHD symptoms in both cord and peripheral blood, and the use of two different technologies (i.e., commercial arrays versus MBD-sequencing). Moreover, as cohorts in the meta-analysis had been designed for different purposes (from general population cohorts to high-risk individuals for psychiatric disorders), ADHD symptom count distributions differed between cohorts; this should positively influence the generalizability of the results of our meta-analysis. Limitations include the use of different reference panels for the deconvolution method for the 450K array versus MBD-seq. While it is important to have platform-specific reference panels, this limited the comparability of data from the two platforms. A second limitation concerned ancestry: the majority of the participants in the included samples were of European ancestry; where this was not the case, ancestry was accounted for by genetic components. In the future, it would be important to include more samples of non-European origin to test the stability of the DNAm-associations across ancestries. Lastly, we might have reduced statistical power as the used reference panels might give different results across age categories, as has been shown for other tissues (84), as well as different ethnicities (85).

In summary, the results of this study show the importance of performing cell type-specific DNAm studies for ADHD (symptoms). We observed cell type-specific associations that were present both in cord and peripheral blood. The differential DNAm patterns for ADHD symptomology were mainly observed in genes involved in signaling through potassium channels and semaphorins. Since this is the first analysis of its kind, replication of the MBD-seq findings and further investigation are warranted. The DNAm at the observed sites may potentially be modifiable. The current findings may thus provide input for studies into ADHD diagnosis and/or intervention.

## Data Availability

All data produced in the present study are available upon reasonable request to the authors.
RaMWAS is freely available from Bioconductor. Scripts used to analyze each of the array data sets are available from GitHub: https://github.com/ejvandenoord/PACE-ADHD-analyses and https://github.com/ejvandenoord/celltype_MWAS

## Acknowledgements

This work was supported by an internal grant from the Donders Centre for Medical Neuroscience of the Radboudumc and by funding for the Dutch National Science Agenda NeurolabNL project (grant 400-17-602). Support was also received from the European Commission’s Horizon 2020 Programme (H2020/2014–2020) under grant agreement n° 728018 (Eat2beNICE). This work reflects only the view of the authors, and the European Commission is not responsible for any use that may be made of the information. This research was supported by the National Institutes of Health (1R01MH104576 to E.J.C.G.v.d.O). In addition, the following personal grants made this work possible: MK was supported by grants from the Netherlands Organisation for Scientific Research (NWO, grant nos. 45219212 and 09150162010073). AH was supported by the HERCULES Center (NIEHS P30ES019776). MCT is funded by a Beatriu de Pinós Postdoctoral Contract awarded by Generalitat de Catalunya-AGAUR and European Commission-Horizon 2020 (2019 BP 00107). BiB is supported by a Wellcome programme grant (WT223601/Z/21/Z)’Born in Bradford: Age of Wonder’ and a joint grant from the UK Medical Research Council (MRC) and UK Economic and Social Science Research Council (ESRC): MR/N024391/1. The work of CAMC, RHM, and JFF is supported by the European Union’s Horizon 2020 Research and Innovation Programme (EarlyCause; grant agreement No 848158). The work of CAMC is supported by the European Research Council (TEMPO; grant agreement No 101039672). CLR. is funded by the MRC (MC_UU_00011/5). DJS and HJZ are supported by the South African Medical Research Council (SAMRC).

A complete overview of all funding sources and acknowledgements can be found in the Supplementary text.

## Availability of data and materials

RaMWAS is freely available from Bioconductor. Scripts used to analyze each of the array data sets are available from GitHub: https://github.com/ejvandenoord/PACE-ADHD-analyses and https://github.com/ejvandenoord/celltype_MWAS

