## Supplementary Text for "Cell type-specific methylome-wide association studies of childhood ADHD symptoms"

Supplementary materials

### Section S1. Methods

#### S1.1. Cohorts

##### S1.1.1 ALSPAC

The Avon Longitudinal Study of Parents and Children is a cohort of 14,541 pregnant women recruited in 1991-1992 in Bristol, UK (1, 2). Live children from these pregnancies have been followed since the start of the study, resulting in a rich dataset of biological, clinical and phenotypic measures from questionnaires and clinics. The study website contains details of all the data that is available through a fully searchable data dictionary and variable search tool (http://www.bristol.ac.uk/alspac/researchers/our-data/). A subsample of these children had DNA methylation measured in cord blood at birth and peripheral whole blood at age 7 within the ARIES project as described in Relton et al., 2015 (3). ADHD was measured via the parent version of the Development and Well-being Assessment (4). The prorated attention/activity score were used in this analysis. All scores were normalized to z-scores. The self-reported ethnicity of all the participants included in this analysis was 'white'. Ethical approval for the study was obtained from the ALSPAC Ethics and Law Committee and the Local Research Ethics Committees. Consent for biological samples has been collected in accordance with the Human Tissue Act (2004). Informed consent for the use of data collected via questionnaires and clinics was obtained from participants following the recommendations of the ALSPAC Ethics and Law Committee at the time.

*Funding*

The UK Medical Research Council and Wellcome (Grant ref: 217065/Z/19/Z) and the University of Bristol provide core support for ALSPAC. This publication is the work of the authors and they will serve as guarantors for the contents of this paper. A comprehensive list of grants funding is available on the

ALSPAC website (http://www.bristol.ac.uk/alspac/external/documents/grant-acknowledgements.pdf). This research was specifically funded by the BBSRC (BBI025751/1 and BB/I025263/1). C.L.R. is funded by the MRC (MC_UU_00011/5).

*Acknowledgments*

We are extremely grateful to all the families who took part in this study, the midwives for their help in recruiting them, and the whole ALSPAC team, which includes interviewers, computer and laboratory technicians, clerical workers, research scientists, volunteers, managers, receptionists and nurses.

##### S1.1.2 Generation R Study

The Generation R Study is a prospective population-based cohort, following children from fetal life onwards (5, 6). Pregnant women residing in the study area of Rotterdam, the Netherlands, with an expected delivery date between April 2002 and January 2006 were invited to participate (N=9,778; participation rate: 61%). The current analyses were limited to children for whom data of the Child Behavior Checklist (CBCL 6-18) at 10 years (7), relevant covariates, as well as DNA methylation at birth (N=1,029) or DNA methylation at 10 years (n=424) were available. DNA methylation was extracted from cord blood at birth and peripheral blood at 10. Bisulfite conversion took place with the EZ-96 DNA methylationTM kit (Zymo, Irvine, CA) and samples were further processed with the Illumina Infinium HumanMethylation450 BeadChip (Illumina Inc., San Diego, CA). Quality control was performed with the CPACOR workflow (8). Arrays with observed technical problems such as failed bisulfite conversion, hybridization or extension, a call rate of =<95% per sample, or sex-mismatches were removed. ADHD was measured with the mother-reported CBCL 6-18 at 10. Seven items were used for the ADHD symptom score, ten were used for the inattention score, and four were used for the hyperactivity score. Each item was rated on a 3-point scale, and items were summed and weighted, allowing for 25%

missing. Last, weighted sum scores were z-transformed. The study has been approved by the Medical Ethical Committee of Erasmus MC, University Medical Center Rotterdam. Written informed consent was obtained for all participating children.

*Funding*

The general design of the Generation R Study is made possible by financial support from the Erasmus MC, Erasmus University Rotterdam, the Netherlands Organization for Health Research and Development and the Ministry of Health, Welfare and Sport. The EWAS data were funded by a grant from the Netherlands Genomics Initiative (NGI)/Netherlands Organisation for Scientific Research (NWO) Netherlands Consortium for Healthy Aging (NCHA; project nr. 050-060-810), by funds from the Genetic Laboratory of the Department of Internal Medicine, Erasmus MC, and by a grant from the National Institute of Child and Human Development (R01HD068437). This project received funding from the European Union's Horizon 2020 research and innovation programme (733206, LIFECYCLE; 848158, EarlyCause; 824989, EUCAN-Connect) and the European Research Council (TEMPO; grant agreement No 101039672).

*Acknowledgements*

The Generation R Study is conducted by Erasmus MC, University Medical Center Rotterdam in close collaboration with the School of Law and Faculty of Social Sciences of the Erasmus University Rotterdam, the Municipal Health Service Rotterdam area, Rotterdam, the Rotterdam Homecare Foundation, Rotterdam and the Stichting Trombosedienst & Artsenlaboratorium Rijnmond (STAR-MDC), Rotterdam. We gratefully acknowledge the contribution of children and parents, general practitioners, hospitals, midwives and pharmacies in Rotterdam. The study protocol was approved by the Medical Ethical Committee of Erasmus MC, Rotterdam. Written informed consent was obtained for all participants. The generation and management of the Illumina 450K methylation array data (EWAS data) for the Generation R Study was executed by the Human Genotyping Facility of the Genetic Laboratory of the Department of Internal Medicine, Erasmus MC, the Netherlands. We thank Mr. Michael Verbiest, Ms. Mila Jhamai, Ms. Sarah Higgins, Mr. Marijn Verkerk and Dr. Lisette Stolk for their help in creating the EWAS database. We thank Dr. A.Teumer for his work on the quality control and normalization scripts.

##### S.1.1.3 INMA

The present study used data from participants recruited between 2003 and 2008 in the de novo cohort sited in Sabadell of the INfancia y Medio Ambiente (INMA) Project, a population-based mother–child cohort study in Spain (9) (study website: http://www.proyectoinma.org/). Cord blood and peripheral blood methylation at 4 years of age was measured using the Infinium® HumanMethylation450 BeadChip. Blood was extracted using the Chemagen kit (Perkin Elmer). DNA concentration was determined by NanoDrop spectrophotometer (Thermo Scientific) and with the Quant-iT PicoGreen dsDNA Assay Kit (Life Technologies). Methylation data was produced in two different laboratories as part of two different projects: in the Genome Analysis Facility of the University Medical Center Groningen (UMCG) in Holland, and in the Bellvitge Biomedical Research Institute (IDIBELL, Barcelona). Both laboratories used the recommended Illumina protocol for the Infinium HumanMethylation450 beadchip. Briefly, 500 ng of DNA was bisulfite-converted using the EZ 96-DNA methylation kit following the manufacturer's standard protocol, and DNA methylation measured using the Illumina Infinium HumanMethylation450 beadchip. DNA methylation data were preprocessed using the minfi package (10). The ADHD symptom score were assessed at the age of 6-9 years using the Attention problems subscale from CBCL syndrome scale as the inattentive score, the Hyperactivity score from Conner's test, and the DSM-ADHD problems score from CBCL DSM scale. All scores were normalized (z-scores). The number of participants with both DNA methylation and phenotype data is 281 for the cord blood study, and 168 for the peripheral blood study. All procedures were approved by the relevant ethics committees, and all parents provided informed consent for their children to participate.

*Funding*

Main funding of the epigenetic studies, and of birth and six to nine years of age assessments/visits in INMA were grants from Instituto de Salud Carlos III (Red INMA G03/176, CB06/02/0041, CP18/00018, PI041436, PI081151 incl. FEDER funds, PI12/01890 incl. FEDER funds, CP13/00054 incl. FEDER funds), CIBERESP, Spanish Ministry of Health (FIS-PI04/1436, FIS-PI08/1151 including FEDER funds, FIS-PI11/00610, FIS-FEDER-PI06/0867, FIS-FEDER-PI03-1615), Spanish Ministry of Economy and Competitiveness (SAF2012-32991 incl. FEDER funds), Agence Nationale de Securite Sanitaire de l'Alimentation de l'Environnement et du Travail (1262C0010), Generalitat de Catalunya-CIRIT 1999SGR 00241, Generalitat de Catalunya-AGAUR (2009 SGR 501, 2014 SGR 822), Fundació La marató de TV3 (090430), EU Commission (261357-MeDALL: Mechanisms of the Development of ALLergy, 308333, 603794, and 634453), and European Research Council (268479-BREATHE: BRain dEvelopment and Air polluTion ultrafine particles in scHool childrEn). We acknowledge support from the Spanish Ministry of Science and Innovation and the State Research Agency through the "Centro de Excelencia Severo Ochoa 2019-2023" Program (CEX2018-000806-S), and support from the Generalitat de Catalunya through the CERCA Program. The work of Marta Cosin Tomas was supported by a Beatriu de Pinós Postdoctoral Contract awarded by Generalitat de Catalunya-AGAUR and European Commission- Horizon 2020 (2019 BP 00107).

*Acknowledgements*

INMA researchers would like to thank all the participants for their generous collaboration. A full roster of the INMA Project Investigators can be found at http://www.proyectoinma.org/presentacion-inma/listado-investigadores/en_listado-investigadores.html.

#### S1.1.4 HELIX

The present study used data from the Human Early Life Exposome Study (HELIX), a collaborative project across six established and ongoing longitudinal population-based birth cohort studies in six European countries (11) (study website: https://www.projecthelix.eu/). For this particular analysis, data from six different HELIX cohorts were used: BIB (United Kingdom), EDEN (France), KANC (Lithuania), INMA (Spain), MOBA (Norway) and RHEA (Greece). From the dataset we included children of European (almost 90%), African, American, East and South Asians, and mixed ancestries (selected by genetic background data). Therefore, models were adjusted for "cohort" and “ethnicity”. DNA was obtained from buffy coat collected in EDTA tubes at age 7-9y. Briefly, DNA was extracted using the Chemagen kit (Perkin Elmer) in batches of 12 samples. Samples were extracted by cohort and following their position in the original boxes. DNA concentration was determined in a NanoDrop 1000 UV-Vis Spectrophotometer (ThermoScientific) and with Quant-iT™ PicoGreen™ dsDNA Assay Kit (Life Technologies). DNA methyaltion was assessed with the Infinium HumanMethylation450 beadchip from Illumina, following manufacturer’s protocol. Briefly, 700 ng of DNA were bisulfite-converted using the EZ 96-DNA methylation kit following the manufacturer’s standard protocol, and DNA methylation measured using the Infinium protocol. A HapMap sample was included in each plate. In addition, 24 HELIX inter-plate duplicates were included. Samples were randomized taking into account cohort, sex and panel. Samples from the panel study (samples of the same subject obtained at two time points) were processed in the same array. Two samples were repeated due to their overall low quality. DNA methylation data were pre-processed using the minfi package (10). The number of participants with both blood DNA methylation (Infinium®HumanMethylation450 BeadChip) and phenotype data (CBCL test) at the age of 7-9 years was 1065. The ADHD symptom scores were assessed at the age of 6-9 years using the Attention problems subscale from CBCL syndrome scale as the inattentive score, the Hyperactivity score derived from Conner's test, and the DSM-ADHD problems score from CBCL DSM scale. All scores were normalized (z-scores). The number of participants with both DNA methylation and phenotype data is 1065. All procedures were approved by the relevant ethics committees, and all parents provided informed consent for their children to participate.

*Funding*

The research leading to these results has received funding from the European Community's Seventh Framework Programme (FP7/2007-206) under grant agreement no 308333—the HELIX project. INMA data collections were supported by grants from the Instituto de Salud Carlos III, CIBERESP, the Conselleria de Sanitat, Generalitat Valenciana, Department of Health of the Basque Government; the Provincial Government of Gipuzkoa, and the Generalitat de Catalunya-CIRIT. KANC was funded by the grant of the Lithuanian Agency for Science Innovation and Technology (6-04-2014_31V-66). The Norwegian Mother, Father and Child Cohort Study is supported by the Norwegian Ministry of Health and Care Services and the Ministry of Education and Research. We are grateful to all the participating families in Norway who take part in this on-going cohort study. The Rhea project was financially supported by European projects, and the Greek Ministry of Health (Program of Prevention of Obesity and Neurodevelopmental Disorders in Preschool Children, in Heraklion district, Crete, Greece: 2011–2014; 'Rhea Plus': Primary Prevention Program of Environmental Risk Factors for Reproductive Health, and Child Health: 2012–2015). The work was also supported by MICINN (MTM2015-68140-R) and Centro Nacional de Genotipado-CEGEN-PRB2-ISCIII. This project has received funding from the European Union's Horizon 2020 research and innovation programme under grant agreement No 874583. The work of Marta Cosin Tomas was supported by a Beatriu de Pinós Postdoctoral Contract awarded by Generalitat de Catalunya-AGAUR and European Commission- Horizon 2020 (2019 BP 00107).

*Acknowledgements*

The authors would like to thank all the participating children, parents, practitioners and researchers in the six countries who took part in this study. The authors would like to thank Sonia Brishoual, Angelique Serre and Michele Grosdenier (Poitiers Biobank, CRB BB-0033-00068, Poitiers, France) for biological sample management and Professor Frederic Millot (Principal Investigator), Elodie Migault, Manuela Boue and Sandy Bertin (Clinical Investigation Center, Inserm CIC1402, CHU de Poitiers, Poitiers, France) for planning and investigational actions. The authors would like to thank Veronique Ferrand-Rigalleau, Céline Leger and Noella Gorry (CHU de Poitiers, Poitiers, France) for administrative assistance (EDEN). The authors would like to thank Silvia Fochs, Nuria Pey, Cecilia Persavente and Susana Gross for field work, sample management and overall management in INMA. The authors would like to thank Georgia Chalkiadaki and Danai Feida for biological sample management, to Eirini Michalaki, Mariza Kampouri, Anny Kyriklaki and Minas Iakovidis for field study performance and to Maria Fasoulaki for administrative assistance (RHEA). The authors would also like to thank Jorunn Evandt, Ingvild Essen for thorough field work, Heidi Marie Nordheim for biological sample management and the MoBa administrative unit (MoBa).

##### S1.1.5 Drankenstein Child Health Study

*Design and study population*

Drakenstein Child Health Study (DCHS). The DCHS, a population-based birth cohort, has been described previously (12). Mothers were enrolled prenatally in their second trimester and followed through pregnancy at two primary care clinics serving two distinct populations (predominantly black African ancestry or predominantly mixed ancestry). Mother-child pairs were followed from birth and infants enrolled in the DCHS were followed until at least eight years of age (12). All births occurred at a single, central facility, Paarl Hospital. The present study is based on children from the DCHS with DNA methylation data from cord blood, genotyping data, and information on ADHD symptoms and covariates.

*Consent and ethical approval*

Ethical approval for human subjects' research was obtained from the Human Research Ethics Committee of the Faculty of Health Sciences of University of Cape Town (HREC UCT REF 401/2009; HREC UCT REF 525/2012). Written informed consent was signed by the mothers on behalf of herself and her infant for participation in this study.

*DNA methylation*

DNA methylation was measured from cord blood collected at delivery by either the MethylationEPIC BeadChips (n=145) or the Illumina Infinium HumanMethylation450 BeadChips (n=103).

*ADHD*

The ADHD symptom score was assessed at an average age of 6.5 years using the Child Behavior Checklist 6-18 (CBCL/6-18), a validated and widely used parental assessment of a child's behavioral and emotional problems (13). Mothers completed questions about a range of emotional and behavioral problems of the child in the past six months on a three-point scale (0=not true, 1=somewhat true, 2=very true). The ADHD symptom score is based on the following CBCL items, which are summarized in an additive score: (1) Fails to finish things, (2) can't concentrate can't pay attention; (3) Can't sit still, restless or overactive; (4) impulsive or acts without thinking; (5) easily distracted; (6) talks too much; (7) is loud. Sub-scores for attention problems are based on items (1), (2) and (5) and hyperactivity/impulsivity problems are based on items (3), (4), (6) and (7). All scores were normalized (z-scores).

*Funding*

The Drakenstein Child Health Study was funded by the Bill & Melinda Gates Foundation (OPP 1017641, OPP1017579), Medical Research Council South Africa, and the National Research Foundation South Africa. Additional support for the DNA methylation work was by the Eunice Kennedy Shriver National Institute of Child Health and Human Development of the National Institutes of Health (NICHD) under Award Number R21HD085849, and the Fogarty International Center (FIC). DJS and HJZ are supported by the South African Medical Research Council (SAMRC).

*Acknowledgements*

The authors thank the study and clinical staff at Paarl Hospital, Mbekweni and TC Newman clinics, as well as the CEO of Paarl Hospital, and the Western Cape Health Department for their support of the study. The authors thank the families and children who participated in this study. The authors also thank Dr. Michael S. Kobor and his team at the University of British Columbia for the generation, pre-processing and quality control of the DNA methylation data (data generation: Julia L MacIsaac, David TS Lin, Katia E Ramadori; pre-processing/quality control: Nicole Gladish).

##### S.1.1.6 Groningen Expert Center for Kids with Obesity

The Groningen Expert Center for Kids with Obesity (GECKO, N=172) was included in the meta-analysis for DNA methylation in cord blood. GECKO is a population-based birth-cohort study of children born in a 1-year period in Drenthe, one of the northern provinces of The Netherlands (14). Children born from April 2006 to April 2007 and living in Drenthe at the time of birth were allowed to participate in this study. The ADHD symptom score was assessed at an average age of 5.9 and 10.7 years using the Strengths and Difficulties Questionnaire (SDQ) questionnaire, and all scores were normalized (z-scores). At birth the obstetricians, midwives or general practitioners took an umbilical cord blood sample that was then processed and stored for future measurements. Written informed consent was obtained from the parents. The study was approved by the Medical Ethics Committee of the University Medical Center Groningen (Medical ethical approval ID: 2005.260).

##### S1.1.7 Lifestyle and environmental factors and their Influence on Newborns Allergy Risk

From the **L**ifestyle and environmental factors and their **I**nfluence on **N**ewborns **A**llergy **R**isk (LiNA) study, we analyzed n=205 10-year-old children whose ADHD symptoms were assessed using the SDQ (Strength and Difficulties Questionnaire). LINA is an ongoing prospective birth cohort study conducted by the Helmholtz Centre for Environmental Research-UFZ in Leipzig, Germany, in collaboration with clinical partners with the aim of linking the impact of environmental factors on the maturation of the immune system in children and the development of allergic diseases, obesity and behavioral disorders. For this study, 629 mother-child pairs were recruited between March 2006 and December 2008 in the city of Leipzig, Germany. Pregnant women were invited to participate and gave informed consent. The study was approved by the ethics committee of the University of Leipzig. Methylation analyses (450K analysis) have been performed in cord blood samples.

*Funding*

This work was supported by an internal grant from the Helmholtz Centre for Environmental Research - UFZ.

##### S1.1.8 POSEIDON

The POSEIDON (Pre, peri, and pOstnatal Stress: Epigenetic Impact on DepressiON) cohort is a longitudinal birth cohort on perceived stress and child development and health (15). 410 mothers were recruited between 2010 and 2013 in the Rhein-Neckar region in Germany. By now, five waves have been conducted during the third trimester of pregnany (T1), at childbirth (T2), 6 months postpartum (T3), at 45 months postpartum (T4) and an online survey during the COVID pandemic (T5). The study protocol was approved by the Ethics Committee of the Medical Faculty Mannheim of the University of Heidelberg. The study was conducted in accordance with the Declaration of Helsinki. Whole cord blood was collected immediately after birth from n=313 newborn singletons. DNA methylation was measured using the Illumina Infinium HumanMethylation450K Beadchip. The ADHD symptom score was assessed as part of the Strengths and Difficulties Questionnaire at the age of 7-9 during the COVID survey.

*Funding*

This work was funded through ERA-NET NEURON "Impact of Early life MetaBolic and psychosocial strEss on susceptibility to mental Disorders; from converging epigenetic signatures to novel targets for therapeutic intervention" [01EW1904]. This work was funded by the Innovationcampus Heidelberg Mannheim Health and Life Sciences (MWK Baden-Württemberg).

*Acknowledgements*

We thank all parents and children for taking part in this study and our student employees and interns for their support with data acquisition and data entry.

##### S1.1.9 Great Smoky Mountain Study

The Great Smoky Mountain Study (GSMS) is a longitudinal cohort of children recruited in eleven contiguous counties in North Carolina, United States of America; this subsample was set up to study the development of child psychiatric disorders (16). Children were included based on a screening questionnaire for parents to identify children with high risk for psychiatric symptoms. The ADHD symptom score was assessed at an average age of 13.5 years using the parent-rated Parent and Child Interview-based Structured Diagnostic Interview (PACA)(16). Subscale scores for IA and HI domains were retrieved, and all scores were normalized (z-scores). In the same assessment, blood spots were collected onto Schleider and Schuell (S&S) filter paper number 903 and long-term stored at -28°C after drying at room temperature and short-term storage (<2 weeks) in a refrigerator. All procedures were approved by the relevant ethics committees, and all parents provided informed consent for their children to participate.

Table S1. Covariates used in methylome-wide association models per cohort

| *Cord blood* | | | | |
|---|---|---|---|---|
| Cohort abbreviation | Demographic covariates | Cell types | PCs on technical probes | PCs on DNA methylation data |
| ALSPAC | Sex, genetic PC1 | CD8T, CD4T cells, NK, B cells, Monocytes, Granulocytes | 7 | 0 |
| GenR | Sex, age | CD8T, CD4T, NK, B cells, Monocytes, Granulocytes | 4 | 5 |
| INMA | Sex | CD8T, CD4T, NK, B cells, Monocytes, Granulocytes | 5 | 2 |
| DCHS | Gestational age, maternal age, age, sex, prenatal smoking, genetic PC1 | CD8T, CD4T, NK, B cells, Monocytes, Granulocytes | 2 | 4 |
| GECKO | Gestational age, maternal age, sex, smoking, ethnicity | CD8T, CD4T, NK, B cells, Monocytes, Granulocytes | 4 | 2 |
| LiNA | Age, sex, smoking, ethnicity | CD8T, CD4T, NK, B cells, Monocytes, Granulocytes, nucleated red blood cells | 2 | 5 |
| POSEIDON | Gestational age, maternal age, age, sex | CD8T, CD4T, NK, B cells, Monocytes, Granulocytes | 12 | 2 |
| *Peripheral blood* | | | | |
| ALSPAC | Sex | CD8T, CD4T, NK, B cells, Monocytes, Granulocytes | 7 | 0 |

| | | | | |
|---|---|---|---|---|
| GenR | Sex, age | CD8T, CD4T, NK, B cells, Monocytes, Granulocytes | 3 | 2 |
| INMA | Sex | CD8T, CD4T, NK, B cells, Monocytes, Granulocytes | 5 | 7 |
| HELIX | Sex, ethnicity, age | CD8T, CD4T, NK, B cells, Monocytes, Granulocytes | 5 | 2 |
| GSMS | Age, age^2, sex, ethnicity, smoking status, socioeconomical status, perceived trauma | T cells, monocytes, granulocytes | 0 | 4 |

For each cohort included in the meta-analysis, individual methylome-wide association studies were run with cohort specific covariates that explained most variation in the data. PC = principal component; mMed = median methylated value; uMed = median unmethylated value

#### S1.2 Quality control, methylation values and technical covariates

##### S1.2.1 Illumina array data

For all cohorts with data from the 450K array quality control was performed according to the data analysis pipeline described previously (1). A user-friendly vignette showing these QC steps and the construction of technical covariates using the RaMWAS (2) software can be found at https://bioconductor.org/packages/devel/bioc/vignettes/ramwas/inst/doc/RW5a_matrix.html. Here we provide a summary.

Quality control (QC):

The minfi R package was used to read raw iDAT files (3). We removed (i) probes in cross-reactive regions, (ii) probes containing a SNP (minor allele frequency > 0.01) within 10 basepairs of the single base extension position (4), (iii) probes with a low bead count (<3 in >1% of the samples) and (iv) probes with a low detection p-value (detection p-value>0.01). Methylation (beta) values were computed by dividing the ratio of the methylated probe intensity by the sum of methylated and unmethylated probe intensities.

Technical covariates:

In addition to demographic and clinical covariates we regressed out lab technical covariates in the MWAS. First, we used the control probes on the array to calculate (i) bisulphite conversion percentages estimated, (ii) median signal intensities for methylated and unmethylated channels, and principal components (PCs) to capture technical variation among individual samples. Second, we calculated indicator variables to regress out effects of the individual BeadChip arrays (slide) and the positional effects on each array (well) which hold 12 samples each. Finally, PCs of the methylation beta values were computed after regressing out all measured covariates to capture any remaining unmeasured confounders.

##### S1.2.2. Methyl-binding domain sequencing (MBD-seq) data

Quality control (QC) and processing of the MBD-seq data was performed using the RaMWAS Bioconductor package(5) (for further details see (6)).

Alignment and QC of reads:

Reads were aligned (build hg19/GRCh37) with Bowtie2(7) using a seed-and-extend approach combined with local alignment while allowing for gaps. Specifically, we used a 20 bp seed with zero mismatches. Rather than considering the entire read, local alignment was used to improve sensitivity by finding the maximum similarity score between the reference sequence and a substring of the extension that may be "trimmed" at both ends. Gaps were allowed to account for small indels. Aligned reads were checked for excessive duplicate reads. That is, if we encountered >3 reads starting at the same location, we assumed these were amplification artefacts and these reads were counted as a single read. The mean number of reads for samples used in this study was 59.7 million (SD=7.4 million) of which, on average, 99% aligned.

QC of samples and CpGs:

Samples were excluded due to (i) failed libraries or sequencing (mainly poor library quality or low number of reads), (ii) poor enrichment and/or high background noise levels, (iii) Sample swap or contamination as determined on the basis of a comparison between GATK(23) SNPs calls from the methylation reads versus GWAS genotypes generated using different blood samples from the same subject(24), (iv) being an (multidimensional) outlier as indicated using the R 'mvoutliers' (https://cran.r-project.org/package=mvoutlier) package that uses principal components of the methylation data as input.

To identify CpGs, we combined reference genome sequence (hg19/GRCh37) with common SNPs calculated on the European super population from 1000 Genomes (Phase 3). To avoid including sites that are CpGs in only a very small proportion of subjects, we excluded CpGs created by SNPs with minor allele frequency <1%. This resulted in 27,916,990 CpGs. CpGs in loci prone to alignment errors, e.g., in repetitive regions, were eliminated prior to the analysis. To identify these CpGs, we used RaMWAS to perform the in silico alignment experiment outlined elsewhere(25) that first creates all possible perfect reads in the reference genome and then aligns these artificial back to the reference. The 365,223 CpGs (1.3%) located in genomic regions showing evidence of alignment problems were removed.

<u>Quantifying methylation and QC of rarely methylated sites:</u>

RaMWAS calculates methylation scores by estimating the number of sequencing fragments covering each of the CpGs(26). With single-end libraries the fragment sizes are not observed. RaMWAS therefore first uses a non-parametric approach to estimate the fragment size distribution from the sequencing data using isolated CpGs. The fragment size distribution is used to calculate the probability that a sequenced fragment will cover the CpG under consideration. For example, this probability is 1.0 for fragments with reads starting within one read-length of the CpG, but is ≤1.0 for fragments with reads starting more than one read-length away. The methylation score is then calculated by taking the sum of probabilities for all fragments aligning within proximity of the CpG and standardized based on the total number of reads. For further details on the interpretation of the methylation score we refer to(27).

Akin to filtering SNPs with low minor allele frequency, we excluded rarely methylated CpGs (average methylation score <0.3). In total 4,881,020 were removed leaving 22,670,747 autosomal CpGs for MWAS, which corresponds to 81% of all common CpGs in the human genome.

<u>Technical covariates</u>:

In addition to demographic and clinical covariates we regressed out lab technical covariates in the MWAS. First, MBD enrichment is affected by the total amount of methylation present on the DNA fragments, which is a function of the number of CpGs and how many of them are methylated. If the amount of MBD protein is low, highly methylated fragments will be overrepresented in the enrichment, and if the amount of protein is too high, non-specific binding will occur(28). Several technical covariates captured sample variation in these enrichment profiles. For examples, we regressed out “peak” which is the CpG density at the point where the methylation score is the highest as well as the number of reads covering sites that were not CpGs. Second, we regressed out the number of reads used to calculate the methylation scores. Finally, PCs of the methylation scores were computed after regressing out all measured covariates to capture any remaining unmeasured confounders.

To identify matching CpG sites between the MBD-seq and 450K array data, we first identified regions consisting of highly correlated methylation marks in the MBD-seq data using an algorithm described elsewhere(18). The average Pearson correlation in these regions, that we label “blocks”, was at least 0.9; MBD-seq CpG sites that were not assayed on the 450K array, could thus still be matched to an array CpG sites if the array site fell in the same block as the MBD-seq CpG site.

#### S1.3 Cell type-specific deconvolution

##### S1.3.1 Intuition behind the method

**Figure S1** provides an intuitive explanation of how the deconvolution works using a hypothetical example involving a deconvolution of bulk blood tissue composed of leukocytes. Since bulk data and proportions of leukocyte A and B will differ between subjects, we can regress bulk levels (Y-axis) on the proportion of leucocyte A (X-axis). The slope of the regression line conveys information about the mean expression levels in leucocyte A and B. For example, extrapolating the regression line to the point where the proportion of leucocyte A is zero (i.e., there are only leucocytes B) estimates the group mean expression in leucocyte B, and extrapolation to the point where the proportion of leucocytes A is one estimates the group mean expression in leucocyte A. By allowing the regression lines to differ between symptom scores, we obtain different predicted cell-type-specific group means that can be tested for significance using standard statistical tests.

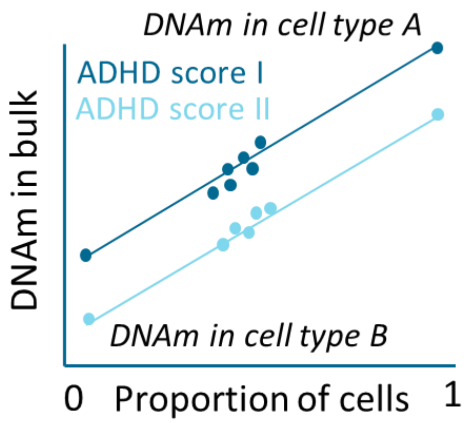


*Figure S1. Deconvolution of cell type-specific effects from bulk data.*

##### S1.3.2 Estimation cell type proportions

To deconvolute cell-type specific effects from bulk data, first cell proportions are estimated for each participant using the bulk DNA methylation data in combination with reference panels that comprise the methylation profiles of purified blood cell-types.

For methylation data from the 450K array, cell type proportions were calculated with *minfi::estimateCellCounts* for either peripheral blood (CD8T cells, CD4T cells, NK cells, B cells, monocytes, and granulocytes) or cord blood (CD8T cells, CD4T cells, NK cells, B cells, monocytes, granulocytes, and nucleated red blood cells)(29). Houseman method(29) was used to estimate the cell type proportions.

For the MBD-seq data, cell type proportion were estimated using a MBD specific reference panel(30) after cell sorting to obtain DNA from the four common cell-types in blood(30): T-cells (CD3+), monocytes (CD14+), granulocytes (CD15+), and B-cells (CD19+). Houseman method(29) was used to generate priors (the estimated means and twice the estimated standard deviations) to obtain final estimates by empirical Bayes using the R 'rstanarm' package (https://cran.r-project.org/package=rstanarm).

##### S1.3.3. Statistical model

The estimated cell type proportions are used to test for association with ADHD symptoms on a cell type-specific level using all study samples with available bulk transcription data. The statistical model we use is (covariates not shown):

$$Y^{bulk} = \sum_{c=1}^{n_c} m_c P_c + \sum_{c=1}^{n_c} m_c^{ADHD}(ADHD \times P_c) + E$$

Thus, measurements from bulk tissue $Y^{bulk}$ are regressed on $c = 1$ to $n_c$, cell type proportions $P_c$, and the product of ADHD symptoms by cell type proportions $(ADHD \times P_c)$. The model allows for covariates and residual effects $E$. Coefficient $m_c$ is the effect of cell type $c$. The symptom count difference $m_c^{ADHD}$ for cell type $c$ is used to test the null hypothesis that cell type means are equal for all symptoms counts. Note that the model has no constant due to $\sum_{c=1}^{n_c} P_c \cong 1$. Alternatively, the model is sometimes written with a constant whereby one of the cell type proportions is omitted (31) but this produces identical results (32-34).

#### S1.4 Pathway and GO analysis

##### S1.4.1 Circular permutations

For the pathway and GO analyses we used circular permutations (35). These permutations account for having correlated test statistics when creating the empirical test statistic distribution under the null hypothesis. When testing if two data sets overlap, we first map the two data sets based on the genomic coordinates of the methylation sites. In the case of a circular permutation, we increase the coordinates of one data set by a randomly chosen number of base-pairs before mapping (we only use the coordinates of the methylation sites to ensure all sites can be mapped). As a result, the overlap between association signals is destroyed (i.e., the “wrong” sites are mapped to each other) but the correlation between the sites in each data set is preserved. For each permutation a different random number is drawn that ranges from a minimum of base-pairs (to avoid true association signal remain) to the total number of base-pairs in the genome. In circular permutations the end of the genome is assumed to be connected to the beginning of the genome (i.e., a “circle”). This is because adding a random number may result in coordinates that are larger than the largest coordinate in the genome. Assuming the genome is a “circle” makes it possible to map such sites again to the beginning of the genome.

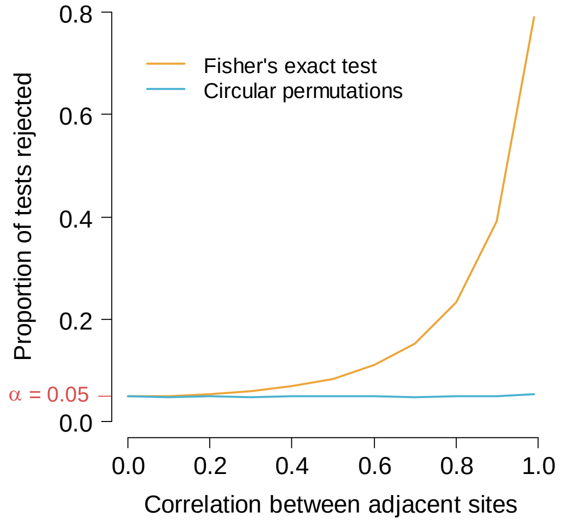


*Figure S1.4.1. Controlling Type I error with highly correlated test statistics*

To demonstrate the validity of this approach, data were simulated assuming the null hypothesis (i.e., no association) was true for 10,000 sites. The correlation between adjacent sites ranged from 0 to 0.99 and decayed as a function of the distance between the sites (i.e., an autoregressive model of order 1 or simplex model). For each chosen value of the site correlation, we performed 10,000 simulations and counted the proportion of tests that were rejected at a P-value threshold of $\alpha$=0.05. Thus, proper control of the Type I error implies that approximately 5% of the tests are rejected.

As indicated above for the permutations we used Cramér's V. For the sake of comparison, the simulated data was also analyzed using Fisher's exact test that assumes independent sites. Figure S2 shows that the use of Fisher's exact test leads to an excess of false positive findings when sites are correlated. For example, when the correlation between adjacent sites is 0.5, there is a two-fold inflation the number of significant tests and this increases to 16-fold inflation if the correlation is 0.99. The use of circular permutations, however, accurately controls the type I error rate even if correlations between adjacent sites are as high as 0.99.

##### S1.4.2 Statistical tests

To study pathways and biological functions, we tested whether top findings were overrepresented in Reactome pathways (36) and Geno Ontology (GO) terms (37), compared to all genes in the human genome. Specifically, we first mapped the MWAS CpGs to genes (ensemble gene annotations grch37, release 91: ftp://ftp.ensembl.org/pub/grch37/release-91/) using the Bioconductor GRanges package. CpGs were allowed to map to multiple genes with flanking regions with size of 100 kB. After mapping we performed 100,000 circular permutations at the CpG level. For each permutation, a two-by-two table was created by cross classifying whether or not the genes were among the top MWAS findings (q-value<0.5) versus whether or not the gene was in the tested pathway. Each gene was counted only once when creating this table (thus, if there were three CpGs in the gene, this was counted as 1 and not as 3). Cramér's V (sometimes referred to as Cramér's phi) was used as the test statistic to measure whether genes from the pathway were overrepresented among the top MWAS genes. P values were calculated as the proportion of permutations that yielded a value equal of greater that Cramér's V observed in the empirical data.

Section 2. Results

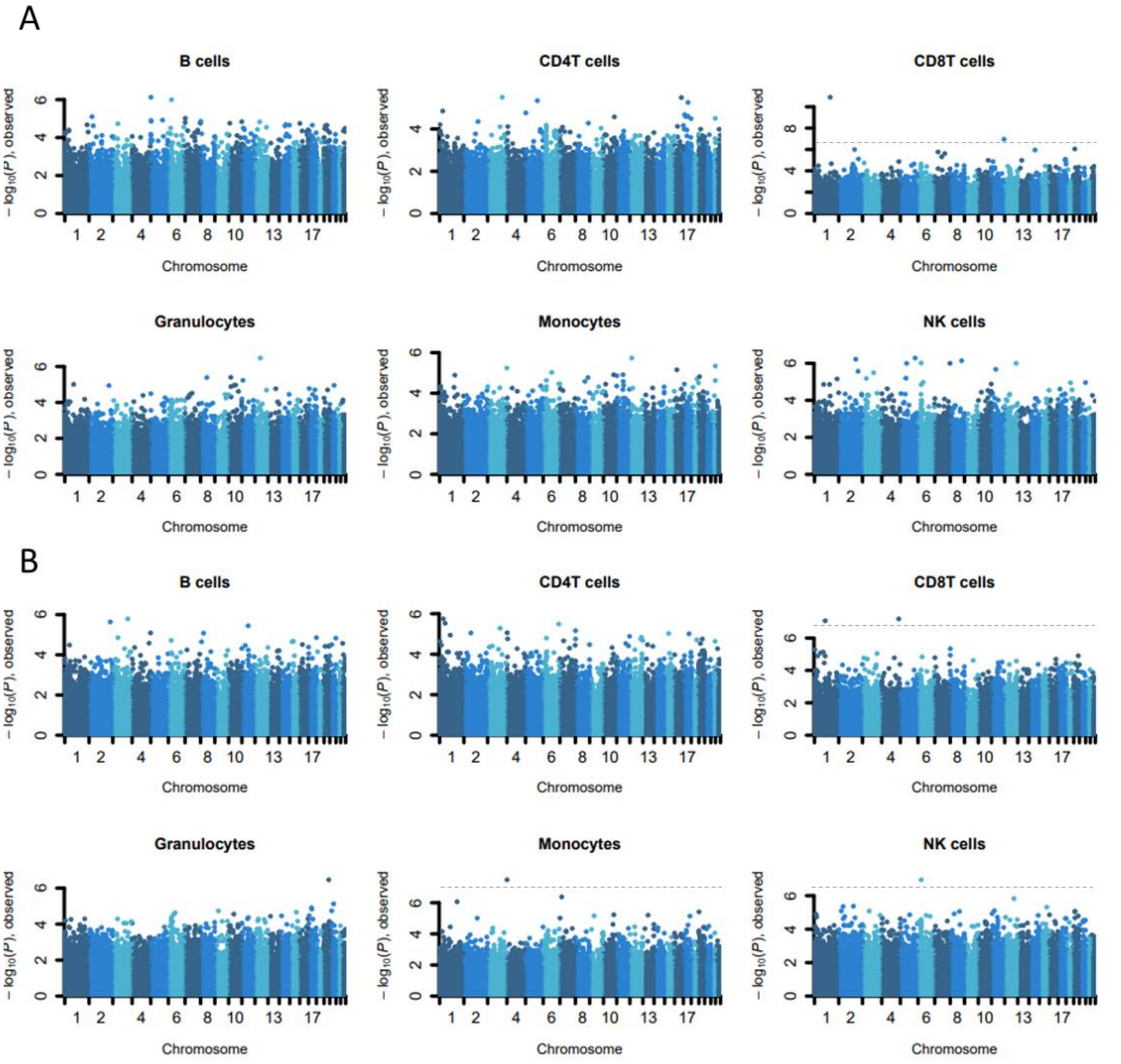


*Figure S2.1. Manhattan plots for methylome-wide association meta-analysis of array-based DNA methylation sites for each cell type in peripheral and cord blood. A) Plots for peripheral blood include DNA methylation data from the GSMS, except for NK cells, since this data was not available. B)*

*Plots for cord blood. The x-axis reflects the chromosomal locations, and the y-axis showed the observed -$log_{10}$(p-value). Dashed grey lines reflect the epigenome-wide significance (q-value<0.05).*

#### Table S2.1 Significant findings of meta-analysis in the individual cohorts

Summary statistics for the epigenome-wide significant sites from the meta-analysis for each cohort separately

Beta= effect size

p-value = unadjusted p-value

df = degrees of freedom

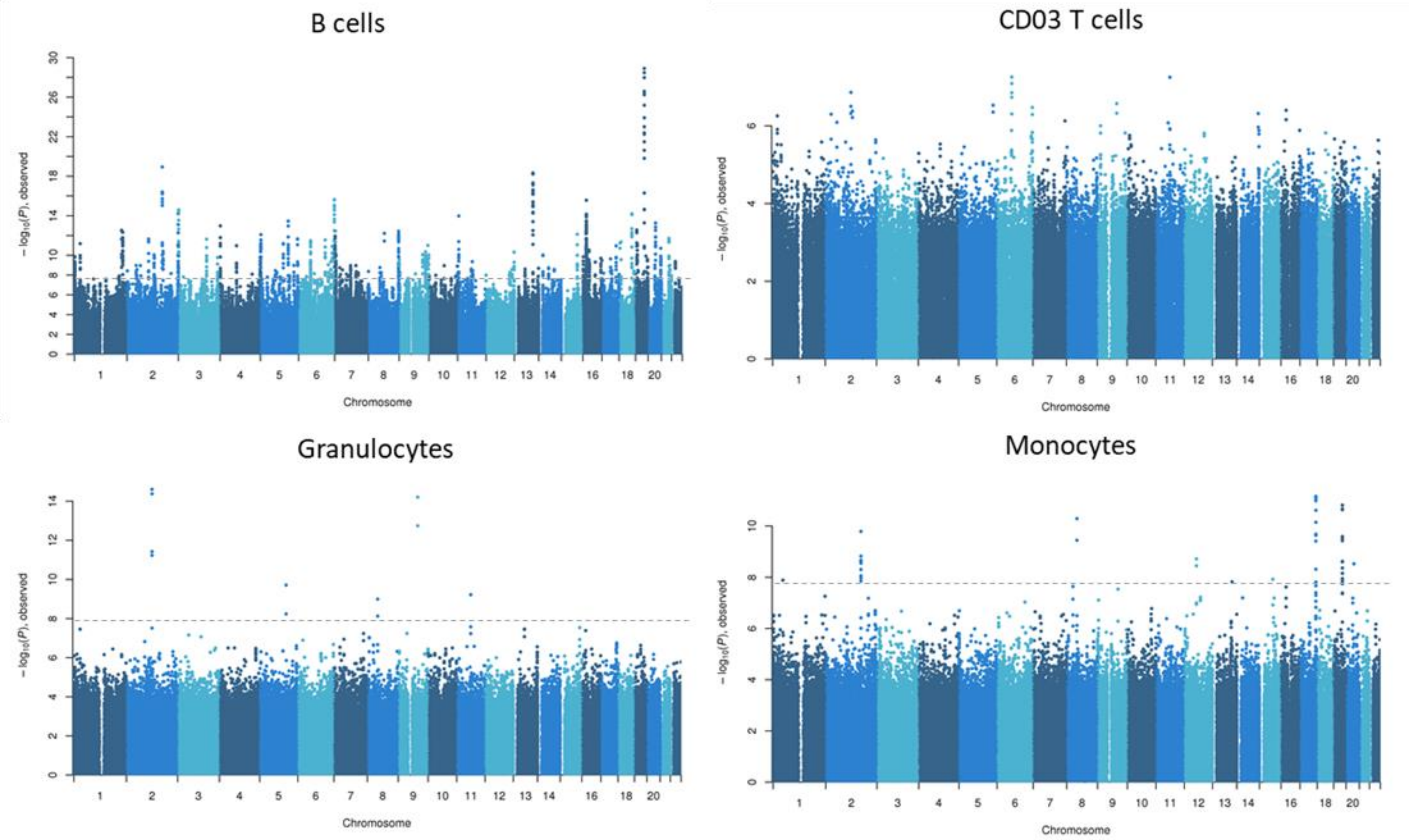


*Figure S2.2.* ***Manhattan plots for methylome-wide association study of MBD-sequencing based DNA methylation sites for each cell type****. The x-axis reflects the chromosomal locations, and the y-axis showed the observed -log10(p-value). Dashed grey lines reflect the epigenome-wide significance (q-value<0.05).*

#### Table S2.2 Significant findings from cell type-specific MWAS on childhood ADHD symptoms in peripheral blood based on MBD-sequencing of 28 million CpG sites

Significant findings for monocytes and granulocytes in GSMS

t-stat = test statistic

p-value = unadjusted p-value

q-value = p-value adjusted for the False Discovery Rate (FDR)

#### Table S2.3 Gene Ontology Term and Reactome enrichment with p<0.05

References supplement

7. Achenbach TMR, L. A. (2000): Manual for the ASEBA Preschool Forms and Profiles. University of Vermont, Research Center for Children, Youth, & Families, Burlington, VT.

13. Achenbach TM, Rescorla LA (2001): Manual for the ASEBA School-Age Forms and Profiles. Burlington, VT: University of Vermont Research Center for Children, Youth, & Families.

21. van den Oord C, Copeland WE, Zhao M, Xie LY, Aberg KA, van den Oord E (2022): DNA methylation signatures of childhood trauma predict psychiatric disorders and other adverse outcomes 17 years after exposure. Mol Psychiatry.
